# Prognostic factors for COVID-19 pneumonia progression to severe symptom based on the earlier clinical features: a retrospective analysis

**DOI:** 10.1101/2020.03.28.20045989

**Authors:** Huang Huang, Shuijiang Cai, Yueping Li, Youxia Li, Yinqiang Fan, Linghua Li, Chunliang Lei, Xiaoping Tang, Fengyu Hu, Feng Li, Xilong Deng

## Abstract

Approximately 15-20% of COVID-19 patients will develop severe pneumonia, about 10 % of which will die if not properly managed. Earlier discrimination of the potential severe patients basing on routine clinical and laboratory changes and commencement of prophylactical management will not only save their lives but also mitigate the otherwise overwhelmed health care burden. In this retrospective investigation, the clinical and laboratory features were collected from 125 COVID-19 patients, who were classified into mild (93 cases) or severe (32 cases) groups according to their clinical outcomes after 3 to 7-days post-admission. The subsequent analysis with single-factor and multivariate logistic regression methods indicated that 17 factors on admission differed significantly between mild and severe groups, but that only comorbid with underlying diseases, increased respiratory rate (>24/min), elevated C-reactive protein (CRP >10mg/liter), and lactate dehydrogenase (LDH >250U/liter), were independently associated with the later disease development. Finally, we evaluated their prognostic values with the receiver operating characteristic curve (ROC) analysis and found that the above four factors could not confidently predict the occurrence of severe pneumonia individually, but that a combination of fast respiratory rate and elevated LDH significantly increased the predictive confidence (AUC= 0.944, sensitivity= 0.941, and specificity= 0.902). A combination consisting of 3- or 4-factors could further increase the prognostic value. Additionally, measurable serum viral RNA post-admission independently predicted the severe illness occurrence. In conclusion, a combination of general clinical characteristics and laboratory tests could provide high confident prognostic value for identifying potential severe COVID-19 pneumonia patients.

**Summary:** With our successful experience of treating COVID-19 patients, we retrospectively found that routine clinical features could reliably predict severe pneumonia development, thus provide quick and affordable references for physicians to save the otherwise fatal patients with the limited medical resource.

## Background

The novel coronavirus (SARS-CoV-2) seems to sweep across the globe ever since its first successful jump from bat to the human being through a yet unknown intermediate(s) approximately in late Nov 2019, still showing a tendency of explosive number increase worldwide [1-3]. The SARS-CoV-2 virus seems more contagious than its sibling virus, severe acute respiratory syndrome (SARS) virus which outbroke in 2003, because over 120,000 individuals contract COVID-19 pneumonia within three months by March 11, which was about 15 times of total SARS cases (8000 in 7 months) [4]. The surging increase of COVID-19 patients within a short time window severely will absorb and occupy the limited medical resource, including physicians, nurses, protective suits, masks, and goggles. Data from the China mainland showed that the majority of total infected patients will recover under simple supervision management, such as quarantined in compartment hospital isolated ward, but that the overall case fatality rate was 2.3% [5]. For the clinical treatment of COVID-19 patients under shortage of enough medical supplies, the critical issue and priority are to treat the severe COVID-19 patients (about 20% of the whole population [5]) and to save their lives with preventive and intensive medical care. However, The clinical presentation of COVID-19 patients differed substantially, including asymptomatic infection, mild upper respiratory tract illness, and severe viral pneumonia[2, 6-8]. Therefore, the most crucial issue is to identify these patients and prioritize their treatment strategy by applying prophylactically medical treatment and management before they progress to the severe stage.

As known, the respiratory function worsens in the severe stage. In the clinical practice, saturated oxygen (< 93% in rest state), reparatory rates (>30 times/min), and deteriorated chest radiology imaging (X-Ray and CT more high resolution) provide references to confirm their severity [5, 9, 10]. Because of the hypoxia stress, most patients will experience an over reactivated immune storm, including elevated their expression level of some specific immunological cytokines and changes of certain types of immune cell counts [6, 11]. Biopsy analysis also showed that the lung bilateral diffuse alveolar damage with cellular fibromyxoid exudates [12]. However, the CT imaging and immunology detection is not only expensive but also far unavailable as for the explosive increase of suspected cases, in particular in those hospitals not well equipped. Can some routine clinical characteristics or/and laboratory measurement, or their combinations predict the occurrence of severe cases?

In this study, we retrospectively analyzed the clinical characteristics of those patients who progressed to severe pneumonia later and found that five simple clinical features and laboratory detection at an earlier time point could serve as prognostic factors facilitating discrimination of severe cases in advance.

## Methods

### Patients

COVID-19 diagnosis was according to the criteria in the new Coronavirus pneumonia diagnosis and treatment plan (trial version 6) issued by the National Health and Health Commission [13]. All 298 COVID-19 patients admitted to Guangzhou Eighth People’s Hospital from January 20 to February 29, 2020, were included in this study. This study complied with the medical ethics of Guangzhou Eighth People’s Hospital. We obtained written consent from the patients.

For this analysis, inclusion criteria were as the following: 1. diagnosed as mild or ordinary on admission; 2. The length of hospitalization > 3 days, and the overall duration of the disease > 7 days. Then, qualified patients were classified into mild symptom group and severe symptom group based on the clinical manifestation. The severe symptom diagnosis was according to criteria as following: 1) Respiratory distress, RR ≥ 30 times/min in the resting state; 2) Oxygen saturation ≤ 93% in the resting state; 3) Arterial blood oxygen partial pressure (PaO2) / oxygen concentration (FiO2) ≤ 300mmHg). The rest of the patients were in the mild group.

### Data collection

Patient general information including gender, age, underlying diseases, epidemic history, etc. and their clinical data including symptoms, signs, clinical classification (course duration> 7 days), laboratory test results and SARS-CoV-2 viral test results were obtained with standardized data collection forms from electronic medical records.

### Statistical analysis

Quantitative data was firstly tested to be normality distribution with the Kolmogorov-Smirnov method. Then, for normalized distributed data, t-test and Tamhane T2 methods were used for variance even and uneven data, respectively. For non-normal data, which was expressed as the median (quartile) [M (P25, P75)], the Mann-Whitney U test was employed. The chi-square test (or Fisher exact probability method) was utilized for analyzing qualitative data. Logistic regression analysis and the receiver operating characteristic curve (ROC) analysis was employed to analyze the independent risk factors. The difference was statistically significant at P <0.05. All analysis was performed using SPSS software (version 20.0).

## Results

### Patient general Information

298 COVID-19 cases (about 85% of total cases in Guangzhou, China) were admitted to Guangzhou Eighth People’s Hospital for treatment from January 20 to February 29, 2020 (Fig. 1). According to the inclusion criteria, 173 cases were excluded for reasons that 23 cases were already in severe symptom stage, 52 cases for a short hospitalization time of < 7 days, and 98 patients for other defects, such as short of a complete set of detection. Finally, 125 cases, including 63 males and 62 females, were qualified to be included for further investigation, and all their disease courses were over seven days, with a maximum of 32 days. Based on the severity of disease at 3-days post-admission, 93 patients fell in the mild group (general 38 cases and mild 55 cases) and 32 patients in severe group (severe 25 cases and critical 7 cases).

**Figure 1.**
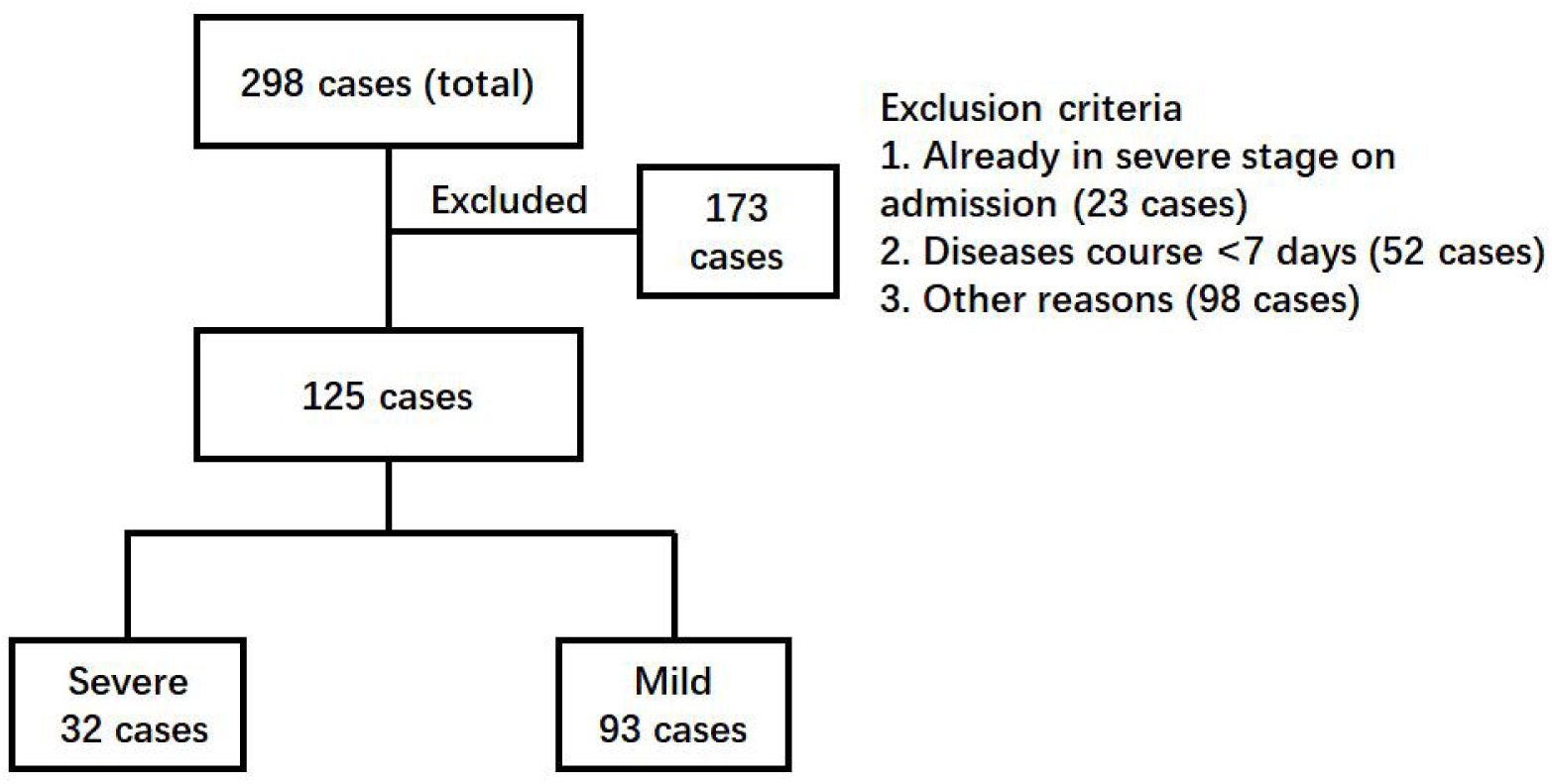
Enrollment chart of COVID-19 patients. The clinical information of a total of 298 patients admitted to the hospital was reviewed. Patients were excluded according to the criteria 1) already in severe stages, 2) with a disease course <7 days, and 3) other reasons such as incomplete detection panel. Finally, 125 patients were further divided into two groups. The severe group included the patients who developed severe COVID-19 pneumonia later (>3 days post-admission). The patients remaining were kept in the mild group.

All included patients aged from 1.5 to 91 years (averaged 44.87 ± 18.55 years) (Table 1). Among them, 37 cases had at least one underlying disease, including 20 cases with hypertension, 8 cases with diabetes, 5 cases with coronary heart disease, 2 cases with chronic obstructive pulmonary disease, 2 cases with chronic kidney disease, 2 cases with chronic liver disease, 2 cases with sleep apnea syndrome. Five individuals with two or more basic disorders and 7 cases with obesity (BMI> 26). Epidemiologically, 88 cases had a history of traveling to or living in the Hubei epidemic area before disease onset. Interestingly, we observed that seven patients developed serum SARS-CoV-2 viral RNA positive after admission but ahead of diagnosis to be a severe symptom.

**Table 1.**
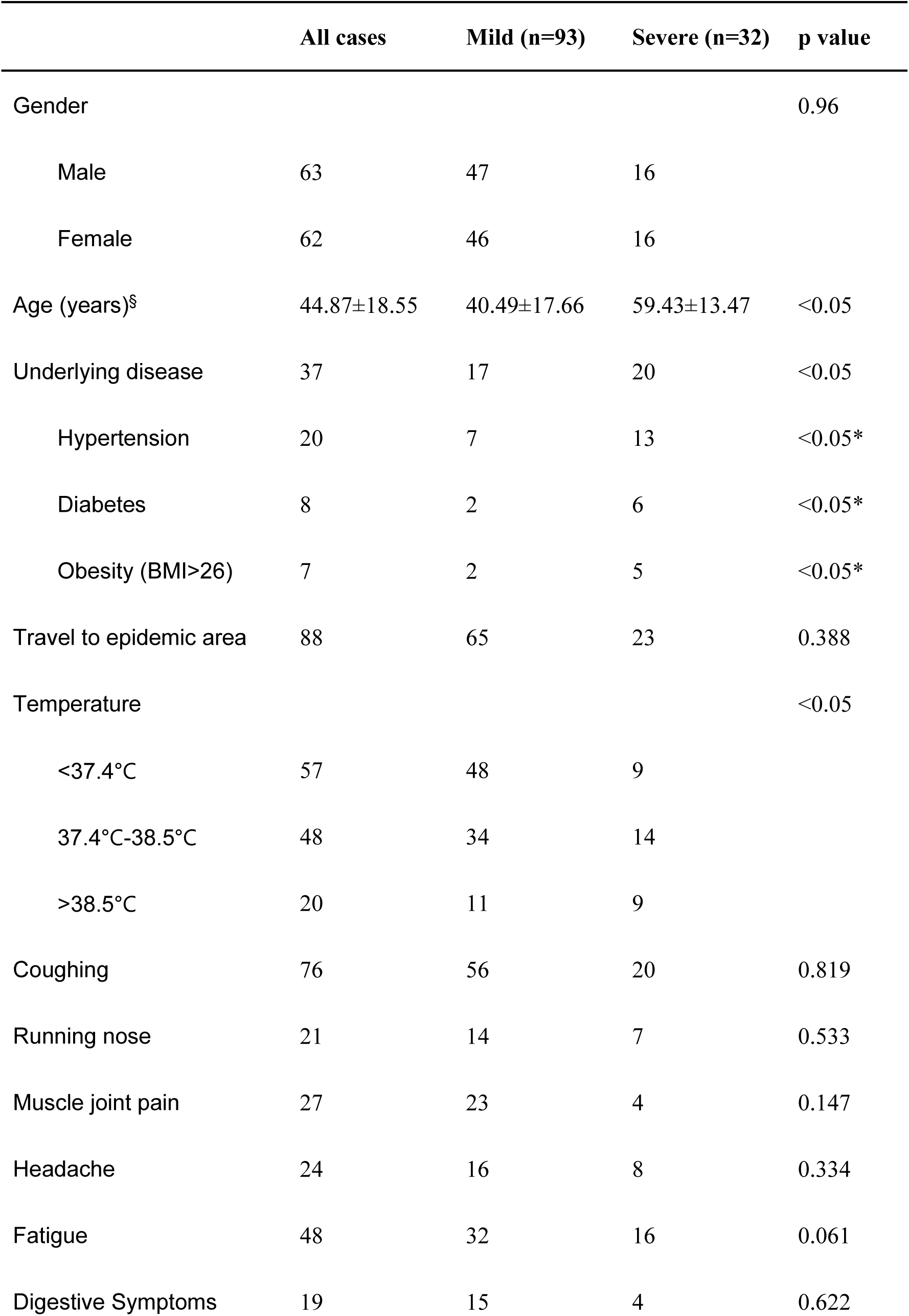
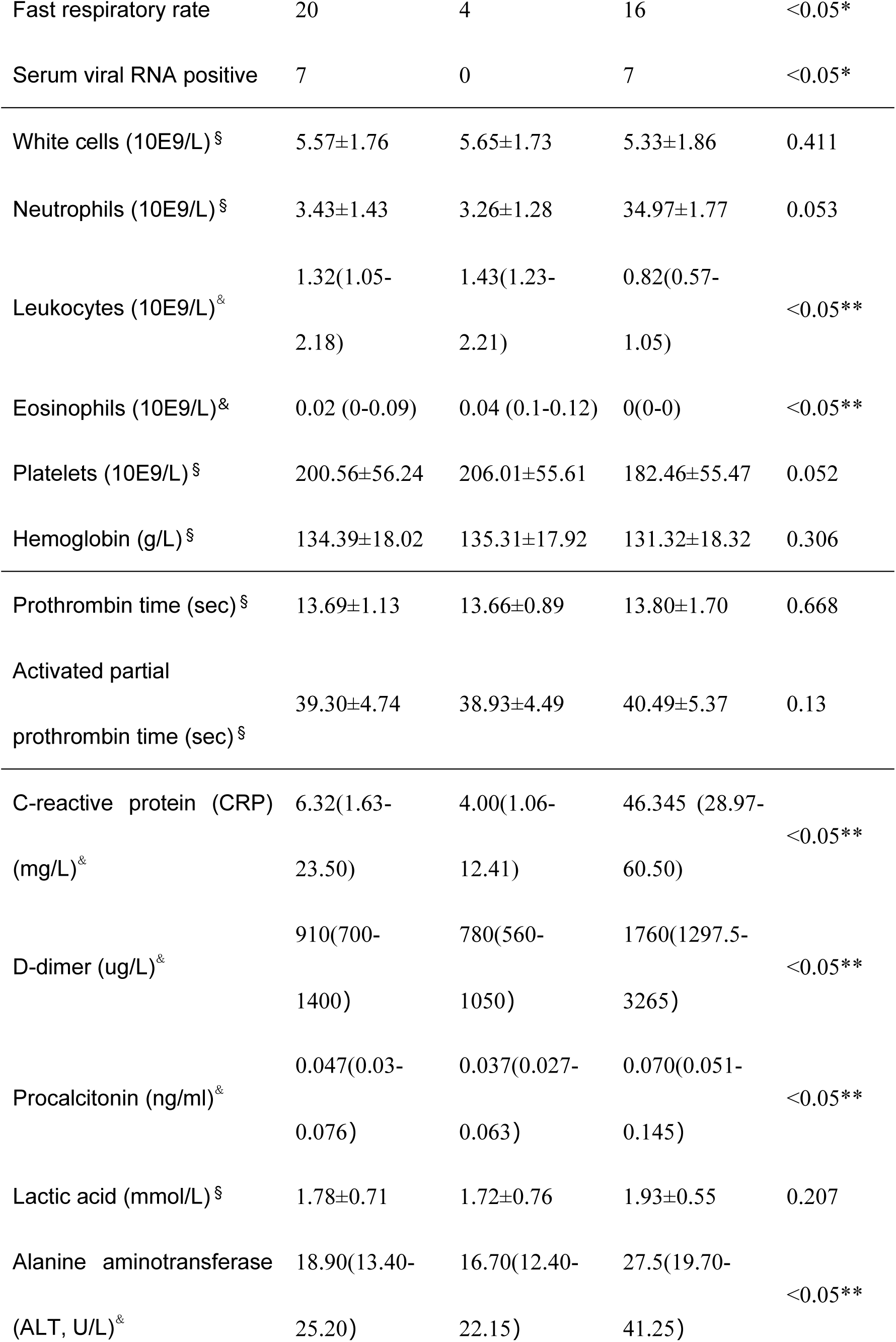
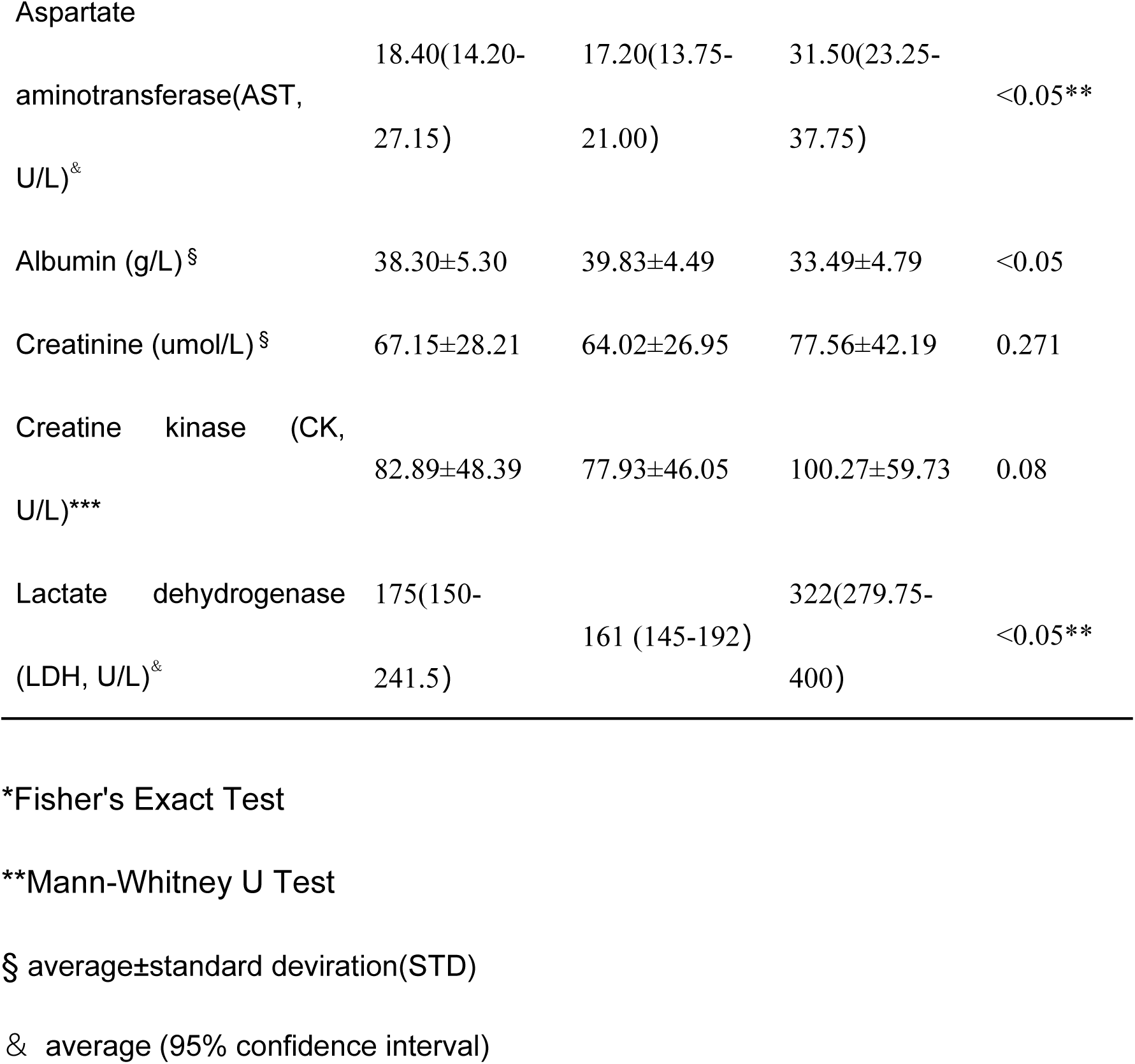
Characteristics of COVID-19 patients

### Factors differed between the mild group and the severe group

The single-factor analysis was applied for each factor between the mild group and the severe group (Table 1). More patients in the severe group were old, with obese (BMI> 26), and with underlying diseases, especially with hypertension and diabetes (P <0.05) compared with the mild group. Among the general factors, no significant difference showed in gender, history of traveling to or living in the epidemic region, coughing, sneezing, muscle joint pain, headache, fatigue, and gastrointestinal symptoms between these two groups (P>0.05). However, more patients in the severe group were with high fever and chest tightness and breath shortness (fast respiratory rate) (P <0.05). The serum concentration of C-reactive protein, procalcitonin, D-dimer, albumin, and lactate dehydrogenase (LDH) increased significantly in the severe group (P <0.05).

Compared to the mild group, patients in the severe group had lower absolute lymphocyte counts and higher eosinophil counts (P <0.05), and similar levels of other parameters, including white blood cells, neutrophils, platelets, hemoglobin, prothrombin time, activated partial thromboplastin time, blood lactic acid, blood creatinine, creatine kinase. Interestingly, the levels of glutamate aminotransferase (ALT) and aspartate aminotransferase (AST) significantly increased for severe patients (P<0.05). However, the median values of ALT and AST were still within the normal range, indicating that most of the severe COVID-19 patients had no significant liver damage.

Importantly, all seven patients with the presence of SARS-CoV-2 viral RNA in blood during the hospitalization, but before being in the severe stage, finally progressed to severe stage, including two severe cases and five critical cases (P <0.05).

### Binary Logistic Regression Analysis of COVID-19 Severe Risk Factors

Next, all categorical variables were converted into covariates, including age, presence of underlying diseases (Yes or No), hypertension (Yes or No), diabetes (Yes or No), obesity (Yes or No), Temperature (<37.4 °C, 37.4-38.5°C, >38.5°C), fast respiratory rate (Yes or No), elevated C-reactive protein (>10 mg/liter), decreased lymphocyte count (<1.1*10E9/L) and eosinophil count (<0.02*10E9/L), elevated procalcitonin (>0.05ng/L), elevated D-dimer (>=2.25 ug/L), decreased albumin (<35g/L), and elevated lactate dehydrogenase (LDH, >250U/L), and subjected to single-factor logistic regression together with multiple independent variables. Those variables with statistical significance were chosen for subsequent binary logistic regression analysis testing the model coefficients, goodness-of-fit, and multicollinearity. Four factors identified to be significantly relevant to the severity of COVID-19 were underlying diseases (X1), fast respiratory rate (>24times/min) (X2), elevated C-reactive protein level (CRP >10mg/L) (X3), and elevated lactate dehydrogenase level (LDH >250U/L) (X4) (Table 2). Finally, the multifactor logistic regression equation was obtained to be logit P = -6.488 + 2.752X1 + 4.056X2 + 2.424X3 + 5.392X4. The β values and odds ratios (OR) for each factor were shown (table 2). The result indicated that elevated LDH ranks the highest correlated with severe symptom development (OR=219.608), followed by the fast respiratory rate (OR=57.726), with underlying diseases (OR=15.67) and elevated CRP (OR=11.289).

**Table 2.** Independent factors associated with severe symptom development in COVID-19 patients

| | Variables | $\beta$ | S.E. | chi-square | P value | OR (95% CI) |
| --- | --- | --- | --- | --- | --- | --- |
| $X_1$ | Underlying diseases | 2.752 | 1.066 | 6.666 | 0.01 | 15.67 (1.94-126.55) |
| $X_2$ | Fast respiratory rate (>24 times/min) | 4.056 | 1.183 | 11.76 | 0.001 | 57.726 (5.685-586.191) |
| $X_3$ | CPR (>10 mg/liter) | 2.424 | 1.004 | 5.823 | 0.016 | 11.289 (1.577-80.838) |
| $X_4$ | LDH (>250 U/liter) | 5.392 | 1.24 | 18.911 | < 0.001 | 219.608 (19.332-2494.742) |
|  | Intercept | -6.488 | 1.499 | 18.738 | 0.001 | 0.002 |

S.E., standard error;
OR (95% CI), Odd Ratio (95% confidence interval).

### The prognostic capacity for severe symptom development

To better evaluate the prediction capacity of each independent risk factors, we plotted their receiver operating characteristic curve (ROC) for the development of severe COVID-19 pneumonia, and calculated their area under the ROC curve (AUC value), sensitivity, specificity, Cut-off value, Youden index and p-value (Table 3). According to the general standard that AUC values between 0.7 and 0.9 means a medium level of diagnostic values, and over 0.9 means a high level of diagnostic values, we observed that all the factors (AUC<0.9) failed to provide high prognostic value when used alone. Then, we two-factor combination test showed that the combination of fast respiratory rate and elevated LDH could provide a high confident prediction (AUC=0.944, sensitivity=0.941, and specificity=0.902) (Table 3). The AUC values of elevated LDH plus underlying diseases or plus elevated CRP were both over 0.9, but their sensitivity or specificity was lower than 0.9. Then, triple factor combination significantly increased the prognostic efficacy, and all combinations had increased sensitivity and specificity (Table 3). Finally, we calculated the prognostic value of the combination of all four factors and found that the AUC value was significantly increased to 0.985 (95% CI 0.968 ∼ 1.000), the sensitivity to 0.912, and the specificity to 0.957 (Table 3).

**Table 3.** Prognostic values for severe COVID-19 pneumonia development.

|  | Factor | AUC (95% CI) | Sensitivity | Specificity | Cut-off value | Youden Index | P value |
| --- | --- | --- | --- | --- | --- | --- | --- |
| Single factor | Underlying diseases (1) | 0.722<br>(0.614~0.829) | 0.618 | 0.826 | 0.367 | 0.444 | <0.001 |
|  | Fast respiratory rate (2) | 0.758<br>(0.648~0.867) | 0.559 | 0.957 | 0.492 | 0.516 | <0.001 |
|  | Elevated CRP (3) | 0.774<br>(0.685~0.864) | 0.853 | 0.696 | 0.298 | 0.549 | <0.001 |
|  | Elevated LDH (4) | 0.855<br>(0.766~0.944) | 0.765 | 0.946 | 0.461 | 0.711 | <0.001 |
| Two factors | (1) + (2) | 0.853<br>(0.767~0.939) | 0.824 | 0.793 | 0.223 | 0.617 | <0.001 |
|  | (1) + (3) | 0.854<br>(0.779~0.928) | 0.853 | 0.696 | 0.274 | 0.549 | <0.001 |
|  | (1) + (4) | 0.940<br>(0.894~0.987) | 0.971 | 0.783 | 0.156 | 0.754 | <0.001 |
|  | (2) + (3) | 0.870<br>(0.795~0.944) | 0.912 | 0.663 | 0.18 | 0.575 | <0.001 |
|  | (2) + (4) | **0.944**<br>**(0.892~0.996)** | **0.941** | **0.902** | **0.315** | **0.843** | **<0.001** |
|  | (3) + (4) | 0.918<br>(0.856~0.981) | 0.765 | 0.946 | 0.365 | 0.711 | <0.001 |
| Three factors | (1) + (2) + (3) | 0.910<br>(0.850~0.969) | 0.765 | 0.902 | 0.253 | 0.667 | <0.001 |
|  | (1) + (2) + (4) | **0.976**<br>**(0.955~0.998)** | **0.912** | **0.935** | **0.411** | **0.846** | **<0.001** |
|  | (1) + (3) + (4) | 0.963<br>(0.933~0.993) | 0.912 | 0.891 | 0.227 | 0.803 | <0.001 |
|  | (2) + (3) + (4) | **0.964**<br>**(0.919~1.000)** | **0.912** | **0.934** | **0.355** | **0.847** | **<0.001** |
| Four factors | (1) + (2) + (3) + (4) | **0.985**<br>**(0.968~1.000)** | **0.912** | **0.957** | **0.374** | **0.869** | **<0.001** |

AUC (95% CI): Area under receiver operating characteristic curve (95% confidence interval).

## Discussion

Our study showed that underlying disease, fast respiratory rate (>24 times/min), elevated serum C-reactive protein level (CRP, >10mg/L), and elevated lactate dehydrogenase level (LDH, >250U/L) were four independent risk factors for predicting the progression of some COVID-19 patients from mild to severe conditions. Firstly, elevated lactate dehydrogenase level ranked as the number 1 (OR=219.332), and fast respiratory rate ranked as the number 2 (OR=57.726) among the four factors (Table 2). Interestingly, elevated lactate dehydrogenase level was associated with severe SARS infection [14], which outbroke in 2003, but was absent in the severe MERS infection[15] which is still circulating. When used individually, all four factors have a moderate prediction value for their low specificity and sensitivity (AUC values <0.9) (Table 3). Secondly, we found that the combination of two factors, fast respiratory rate plus elevated LDH, could provide a high prognostic value for severe symptom development (AUC=0.944, sensitivity=0.941, and specificity=0.902). Combinations of triple factors could significantly increase the prognostic value (AUC>0.9). Finally, a combination of all four factors, provide an excellent prognostic efficacy, achieving AUC=0.985 (95% CI 0.968 ∼ 1.000) with high sensitivity (0.953), and specificity (0.968).

Our hospital has treated over 80% of COVID-19 patients in Guangzhou city, 298 cases as of February 29, 2020, including 55 severe cases, but only one death case. All the patients except two patients recovered as of March 15. A retrospective analysis of all the cases revealed that the extremely low fatality rate in our hospital (1 of 298 cases, 0.0336%, which was pretty lower than the overall fatality rate (2.3%) in China [5]) was largely attributed to the effect of an expert panel, consisting of physicians from multiple disciplines, including infectious diseases, respiratory diseases, and intensive care unit (ICU), and radiology. Patients newly admitted were reviewed by the panel, and patients who meet several of the following criteria were transferred immediately to the ICU isolation ward for close supervision, including, (1) the illness onset has entered 7-10 days; (2) over 50 years old; (3) obesity, pregnant women, children; (4) with underlying diseases, especially hypertension, diabetes, COPD; (5) fast respiratory rate ; (6) obvious decline in spirit and appetite; (7) Significant reduction and/or progressive decline of peripheral blood lymphocytes; (8) Decreased in albumin; (9) elevated C-reactive protein (10) elevated lactate dehydrogenase; and (11) quickly deteriorated, or with two or more lesions in lungs revealed by chest imaging. Once they progressed to the severe stage, they received treatment immediately. The above four prognostic factors, as routine and affordable clinical characteristics, were included in these criteria and facilitated their immediate and preventive therapy from a retrospective aspect.

All the seven patients who were detected to be serum viral RNA positive developed severe symptoms very soon, which further confirmed our previous observation that detectable 2019-nCoV viral RNA in Blood is a reliable indicator for the further clinical severity [16]. However, as the viral RNA positive rate were low high (7cases of 32 cases, 21.8%) in this study and from other reports [17] and viral RNA detection is expensive, we do not recommend to continuously detecting viral RNA. In this regard, we suggest reserving the precious reagent for confirming virus infection.

In conclusion, our study indicated that underlying disease, fast respiratory rate, elevated serum C-reactive protein level, and elevated lactate dehydrogenase level significantly correlated to the development of severe COVID-19 pneumonia, and that elevated lactate dehydrogenase and fast respiratory rate or plus one or two more other factors can serve as prognostic factors for the discriminating potential severe cases among the mild COVID-19 patients. Our study provided convenient, reliable, and affordable references for both patients and physicians to make a high confident decision to commence management and treatment safely.

## Data Availability

This is a retrospective analysis of clinical data.

## NOTES

### Contributors

Huang Huang, Feng Li, and Xilong Deng conceived the study, and wrote the manuscript. Huang Huang, Yongjiang Cai and collected data and performed data analysis. Huang Huang, Yongjiang Cai, Yueping Li, Youxia Li, Yinqiang Fan, and Xilong Deng participated in the clinical treatment. Linghua Li, Chunliang Lei, and Xiaoping Tang supervised the clinical treatment. Fengyu Hu analyzed the results. All authors read the manuscript and approved the final version.

#### Acknowledgments

The authors would like to thank all nurses in the treatment team for taking care of the patients and doctors in the expert panel for their treatment guidance.

